# Long-term Services and Supports, Cognitive Impairment, and Disease Management among Older Chinese Adults

**DOI:** 10.1101/2021.09.10.21263411

**Authors:** Zhuoer Lin, Xi Chen

**Affiliations:** Department of Health Policy and Management, Yale School of Public Health; Department of Economics, Yale University; Alzheimer’s Disease Research Center, Yale University

## Abstract

Rapid population aging elevates burden of chronic and non-communicable diseases among older adults. Despite the critical role of self-management in disease prevention and control, effective management of diseases can be cognitively demanding and may require additional supports from family, friends and social services. Using nationally representative data from China, this paper documents the gradient relationship between cognitive impairment and disease management, and characterizes the differential effects of long-term care services and supports (LTSS) on disease management among older adults in different stages of cognitive impairment. In specific, we examine preventive care use and the management of hypertension, a highly prevalent but inadequately addressed chronic disease in China. We find that a severer stage of cognitive impairment is associated with poorer performance in disease prevention, hypertension awareness and management. While some of the LTSS offered by spouse, friends or community significantly facilitate active disease management behaviors, the effects are only evident among older adults with no cognitive impairment. By contrast, we find no significant effect of LTSS among cognitively impaired individuals. These findings reveal the vulnerability of older adults with cognitive impairment in disease management, and point to the importance of promoting targeted interventions to reduce barriers of receiving and utilizing LTSS, especially among cognitively impaired population.

## 1. Introduction

The world population is aging rapidly with a rising burden of chronic and non-communicable diseases (NCDs) (Bennett et al., 2018). Faced with enormous challenges in supporting older population, disease prevention and control are of crucial importance for individuals, families, and society. However, the performance of chronic disease management is far from satisfactory. The overall diagnosis, treatment and control rates of hypertension, for instance, are low in some developed countries; and the rates are even worse in developing countries, including China (World Health Organization, 2019). Moreover, as the population ages, a growing size of population are living with cognitive impairment (CI) or even dementia, which makes disease management even more difficult and demanding (Livingston et al., 2020; Winblad et al., 2016). Declining cognitive function may interfere with patients’ ability to detect health changes and adhere to disease management strategies, engendering new barriers for effective disease management and control, thereby increasing the risk of CI, leading to a vicious cycle (Feil et al., 2012).

The challenges are particularly acute in China, where a huge number of older adults are living with NCDs, CI and dementia (Chan et al., 2013; Jia et al., 2020; Yang et al., 2013; Zhou et al., 2019, 2016). While long-term services and supports (LTSS) are expected to play pivotal roles in China amid the rapid aging trend, the capacity of the system that cares for older adults is still concerning. Notably, with reduced fertility in the past decades, coupled with rapid urbanization and increased labor mobility, informal caregiving and support provided by family members, relatives and friends is being increasingly strained (Feng et al., 2020, 2012). Meanwhile, despite recent efforts to reform long-term care system, formal LTSS, such as home and community-based services (HCBS), are still underdeveloped (Feng et al., 2020; Wang et al., 2018). Facing the marked demographic shifts and system changes, it remains unclear whether the current informal and formal LTSS in China are effective in facilitating disease prevention and control, especially among cognitively impaired individuals with increased difficulties in obtaining services and supports.

Using nationally representative physical examination and survey data from the China Health and Retirement Longitudinal Study (CHARLS), this paper assesses the availability of LTSS among Chinese older adults and evaluates their impacts on preventive care utilization, hypertension awareness and management. In particular, we investigate the heterogeneity across older adults in different stages of CI.

We find that, LTSS are generally inadequate among Chinese older adults, particularly among those with CI. The overall utilization rates of home- and community-based services are quite low among all older adults. Older adults with severer stages of CI are less likely to have spousal and friends’ supports. The performance of disease management is consistently poorer among cognitive impaired older adults; and the gradient relationship is corroborated by our regression analysis when adjusting for individual sociodemographic attributes and geographical differences. Specifically, we find older adults with mild CI or severe CI have lower chance of receiving physical examination, become less aware of own hypertension status, and tend not to monitor blood pressure (BP) regularly or even annually, as compared to those without CI. Specifically, severe CI, for example, leads to 38% lower odds of annual physical examination, 44% lower odds of annual BP examination, 57% lower odds of regular BP examination, and 66% lower odds of hypertension awareness relative to no CI.

Access to LTSS benefits older adults’ disease management, with informal supports being more impactful than formal services. In particular, the effects of spousal and friends’ supports are large and significant for disease prevention or BP management; whereas formal care, such as community-based services, only have marginal effect on regular BP examination. Moreover, we demonstrate that the effects of LTSS differ for older adults with and without CI. Some LTSS, such as spousal and friends’ supports, only have strong and significant impact for cognitively normal individuals. By contrast, no significant effect of LTSS on disease management are found for cognitively impaired individuals, though there is some modest evidence suggesting the potential role of home-based and community-based services in equalizing the gaps across individuals in different stages of CI.

These findings may contribute to the existing literature in several ways. First of all, we are one of the first to document the cognitive gradients in a broad range of disease management outcomes, both in prevention care utilization and hypertension management. We also show differences in the availability of LTSS. Previous studies largely focus on the impact of cognitive ability on the self-management behaviors of a particular disease, usually with small sample size (Feil et al., 2012; Hajduk et al., 2013; Kiza and Cong, 2021; Lovell et al., 2019; Sinclair et al., 2000). Less attention is paid to disease prevention. Studies in China, on the other hand, mostly focus on the socioeconomic differences in disease management, neglecting the potential heterogeneity by cognitive ability (Feng et al., 2014; Li and Lumey, 2019; Lu et al., 2017; Zhao et al., 2016). Our study thus fills the gap by highlighting the high vulnerability of cognitively impaired individuals in utilizing preventive care, and managing chronic disease and condition.

Second, we provide novel evidence on the impacts of informal and formal LTSS on the performance of disease management. While there is a growing body of studies showing the health impacts of LTSS, such as their impacts on physical function, mental health, harms, and mortality, evidence on disease prevention or chronic disease management is limited (Sohn et al., 2020; Wysocki et al., 2015). In this study, we enrich our understanding of the effects of LTSS on both preventive care utilization and the management of hypertension among Chinese older adults. Our results suggest that informal care still plays a critical role, while formal care only has modest impact on disease management. Continuous efforts, therefore, should be devoted to developing accessible, high-quality, and well-targeted home- and community-based services.

Finally, we reveal the differential effects of LTSS on disease management among older adults in different stages of CI. To our best knowledge, no study to date has shown the heterogenous effect of LTSS on disease management by stage of CI. In our study, we demonstrate that cognitively impaired older adults in China tend to benefit less from LTSS compared to those without CI. Our findings thus emphasize the increased vulnerability of older adults with CI in disease management, especially with weakening informal care, and point to the importance of promoting targeted interventions to reduce their barriers of receiving and utilizing LTSS.

## 2. Methods

### 2.1 Data and Participants

Data are obtained from China Health and Retirement Longitudinal Study (CHARLS), a nationally representative survey of Chinese older adults age 45 years or older. The national baseline was conducted in 2011, and the respondents were longitudinally followed up in 2013, 2015, and 2018. In each wave, comprehensive data on participants’ demographic characteristics, family, health status and health care utilization, cognition, and work and economic conditions were collected (Zhao et al., 2014). Additionally, physical measurements, such as BP, were taken in 2011, 2013, and 2015. Each participant’s systolic BP and diastolic BP were recorded three times by a trained nurse in 2015, and his or her medication usages were interviewed, which can be used to assess hypertension status. Yet, these biomarkers are not available for wave 2018 due to a revised survey design (Zhao et al., 2020).

In the first 3 waves, cognitive ability is assessed with a reduced form of the Telephone Interview for Cognitive Status (TICS) as well as the HRS version of CERAD immediate and delayed word recall. Starting in 2018, CHARLS undertook an expanded set of cognitive tests for older adults age 60 years and over, which includes Mini-Mental State Exam (MMSE), brief community screening instrument for dementia CSI-D, and other cognitive instruments (Zhao et al., 2020). These tests are well established and have been widely used to assess individuals’ status of CI and dementia (Borenstein and Mortimer, 2016). Besides, as compared to previous three waves, 2018 survey additionally collects data on long-term care utilization, thereby allowing us to examine long-term services and supports.

Our sample are constructed primarily based on CHARLS 2018 survey as it has comprehensive cognitive assessment and detailed long-term care measures. Sample inclusion criteria are illustrated in Figure 1. We first include 3,471 participants with valid measures of cognitive assessment (i.e., MMSE), LTSS, and basic individual sociodemographic characteristics from 2018 survey. These sample are used to examine the effects of LTSS and cognition on preventive care utilization. Then, we identify sample that has underlying risks of hypertension based on biomarkers, i.e., BP, collected in 2015, to assess the performance of hypertension management. A total of 1,364 participants are identified as hypertensive based on the biomarkers, and are used to examine their awareness of the condition as of 2018 survey^1^. To investigate health education and monitoring of hypertension, we further include older adults who reported diagnosed hypertension in 2018 survey, along with the sample identified based on biomarkers, which leads to 1,741 participants.

**Figure 1.**
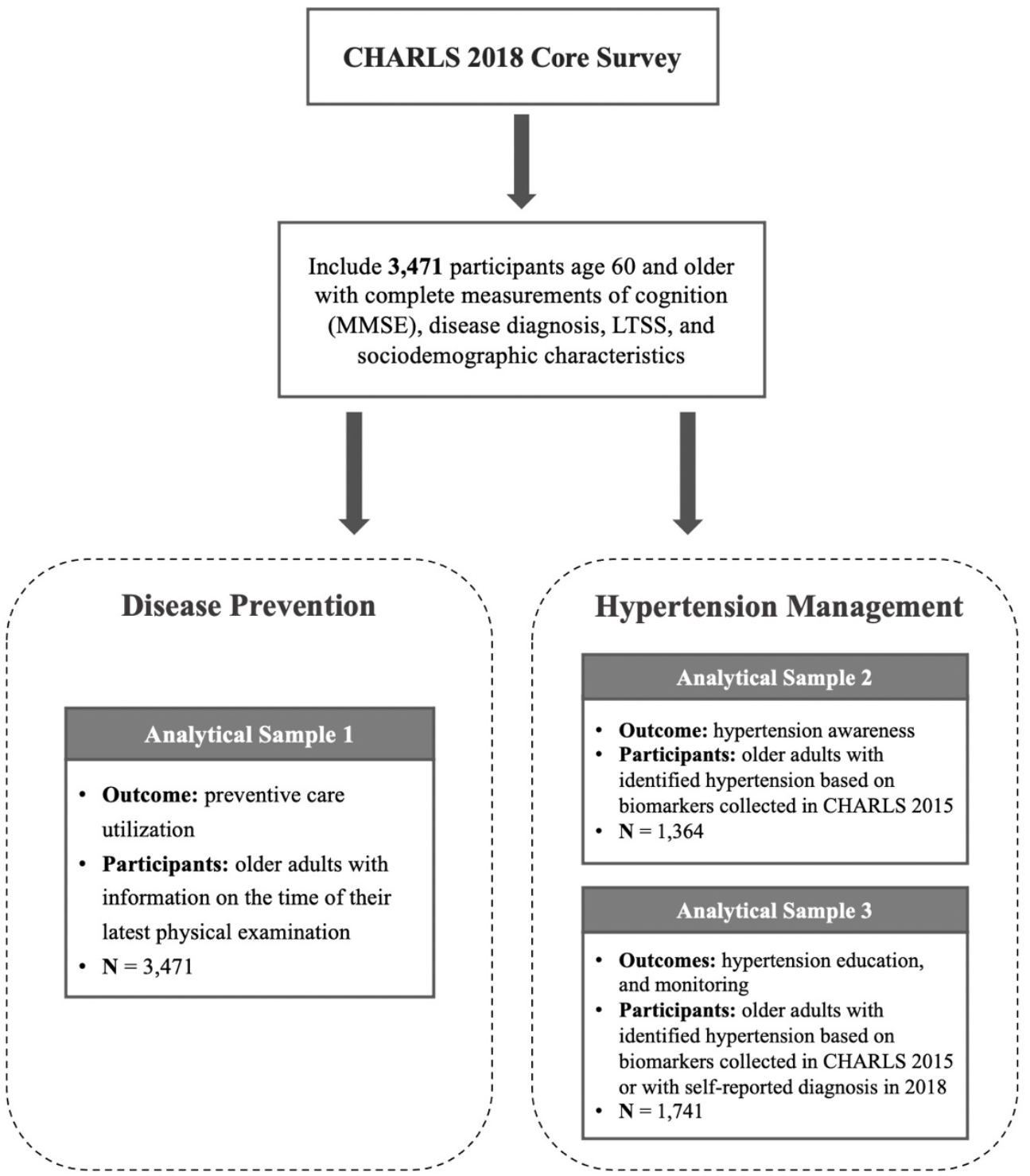
Flow chart of sample inclusion and study participants *Notes:* CHARLS = China Health and Retirement Longitudinal Study; MMSE = Mini-Mental States Examination; LTSS = Long-term Services and Supports. Biomarkers, including systolic and diastolic blood pressure, were collected by a trained nurse in CHARLS 2015 wave.

### 2.2 Cognitive Status

Cognition of older adults is assessed with the Mini-Mental State Examination (MMSE), a 30-score scale that is designed to evaluate major domains of cognitive status, including orientation, attention, calculation, memory and language abilities (Folstein et al., 1975). As a validated cognitive instrument, the MMSE is widely used in clinical practice to screen for CI and dementia. According to an established classification criteria, participants are classified with the following cut-off scores: no CI, 24-30; mild CI, 18-23; and severe CI, 0-17 (Tombaugh and McIntyre, 1992). The MMSE is only measured among older adults age 60 years and over in CHARLS. The distribution of MMSE for our sample is illustrated in Figure A1.

### 2.3 Long term Services and Supports

We measure both access to informal LTSS and formal LTSS in our analysis. Informal LTSS represent the support that is provided by informal caregivers, such as children, spouse, friends and other relatives. In this study, we measure the participants’ supports received from their children, spouse, and friends. Children’s support is assessed by adding up the total number of days the respondents have in-person contact with each children. Individuals whose children visiting them more often than sample median are considered to have good access to children’s support. Spousal support is measured by individuals’ marital and co-residence status. Individuals who were married and living with spouses are considered having spousal support. Lastly, friends’ support is assessed by the social networks and support individual possess. Participants were asked to report whether they had interaction with their friends in the past month. Individuals who had social interaction are considered to have friends’ support.

Formal LTSS reflect the support and care obtained through professional agencies. In China, formal LTSS have two key components, including home-based services and community-based services. In this study, we consider services that can be clearly classified into these two categories. For home-based services, we include the onsite visits and family beds; while for community-based services, we include day care centers and community nursing. Participants are asked to report if they have ever received each service^2^. Older adults are considered to have good access to the services if any participants in their local city have ever used these services^3^. We also evaluate the access at the community-level as a robustness check, and the results are robust to this specification.

### 2.4 Disease Management

In this study, we focus on the utilization of preventive care, as well as the management of hypertension, a highly prevalent but inadequately controlled chronic disease in China. Disease prevention is measured by preventive care utilization. Participants were asked to report the time of their last physical examination (CHARLS physical examination in 2015 excluded). According to the reported exam date and interview date, we evaluate whether the participants took any physical examination within 1 year, within 3 years, or not. This is intended to measure the extent to which individuals actively monitor their overall health conditions, regardless of their illnesses.

The performance of hypertension management is evaluated based on hypertension awareness, health education, and monitoring. Participant’s hypertension status is determined primarily based on biomarkers collected in 2015. An individual is considered hypertensive if he or she had an average systolic blood pressure (SBP) of ≥ 140 mm Hg, an average diastolic blood pressure (DBP) of ≥ 90 mm Hg, or self-reported use of antihypertensive medication (Feng et al., 2014; Lu et al., 2017). Participants with identified hypertension are deemed aware of their conditions if they reported they had ever been diagnosed by a doctor by 2018. Besides, in 2018 survey, participants were asked if they have ever received any medical advice or health education regarding hypertension management, including weight control, exercise, diet, smoking control; and how they monitored their BP in the past 12 months. Specifically, participants were asked if they had at least one BP test annually, and if they had regular BP examination by a doctor. Participants who reported hypertension diagnosis in 2018, in addition to the hypertensive cases identified in 2015, are included to investigate their overall performance of hypertension education and monitoring.

### 2.5 Statistical Analysis

As descriptive analyses, we examine the differences in sociodemographic characteristics, access to LTSS, and disease management across older adults in different stages of CI (i.e., no CI, mild CI, and severe CI). Chi-square test is employed for categorical variables and one-way ANOVA is employed for continuous variables.

To estimate the effects of CI and LTSS on various binary disease management outcomes, logistic regressions are applied in this study. The model can be specified as,

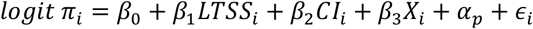

where π_*i*_ represents the probability of disease management outcome *Y*_*i*_ = 1 for individual *i. LTSS*_*i*_ are the primary variables of interest, which consists of a series of dummy variables indicating whether individuals have access to informal care (i.e., children, spouse, and friends’ support) and formal care (i.e., home-based services, and community-based services). *CI*_*i*_ denote the cognitive status of the individual, which include two dummies indicating whether the individual have mild CI or severe CI. No CI is considered as the reference group. *X*_*i*_ are a set of individual sociodemographic attributes measured as of 2018 survey, including age, gender, years of schooling, rural/urban hukou status, number of chronic diseases, activities of daily living (ADL), instrumental activities of daily living (IADL), number of living children, and health insurance^4^. These variables are included to control for heterogeneity in individual characteristics. Besides, province fixed-effects, *α*_*p*_, are included to account for time-invariant geographical differences across provinces.

Moreover, to explore the heterogeneous effects, we stratify our sample by cognitive status and estimate the models respectively for individuals with and without CI. All the analyses are performed in Stata 17.0. Standard errors are clustered at city level.

## 3. Results

### 3.1 Descriptive Findings

Table 1 shows the sociodemographic characteristics of our sample. Of the 3,471 older adults included, 2,440 (70.3%) have no CI, 799 (23.0%) have mild CI, and 232 (6.7%) have severe CI as measured in 2018. Older adults who are female, older aged, higher educated, those who have rural hukou and rural medical insurance, and those who have more living children and chronic diseases are more likely to be in severer stages of CI.

**Table 1.** Characteristics of older adults with different stages of cognitive impairment (CI)

|  | (1)<br>No CI<br>(N=2440) | (2)<br>Mild CI<br>(N=799) | (3)<br>Severe CI<br>(N=232) | (4)<br>*p*-value |
| --- | --- | --- | --- | --- |
|  | Mean<br>(SD) | Mean<br>(SD) | Mean<br>(SD) |  |
| Female | 0.330<br>(0.470) | 0.344<br>(0.475) | 0.513<br>(0.501) | <0.001 |
| Age | 67.56<br>(5.702) | 68.23<br>(6.321) | 70.39<br>(7.260) | <0.001 |
| Years of Schooling | 7.950<br>(3.971) | 5.577<br>(4.128) | 2.211<br>(3.413) | <0.001 |
| Rural/Urban Status | 0.543<br>(0.498) | 0.735<br>(0.442) | 0.901<br>(0.299) | <0.001 |
| Number of Chronic Diseases | 2.496<br>(1.906) | 2.632<br>(2.088) | 2.487<br>(2.019) | 0.235 |
| IADL (0-5) | 0.182<br>(0.575) | 0.394<br>(0.913) | 0.763<br>(1.227) | <0.001 |
| ADL (0-5) | 0.194<br>(0.615) | 0.407<br>(0.950) | 0.595<br>(1.180) | <0.001 |
| Number of Living Children | 2.719<br>(1.476) | 3.056<br>(1.477) | 3.716<br>(1.772) | <0.001 |
| Urban employee MI | 0.346<br>(0.476) | 0.164<br>(0.370) | 0.065<br>(0.246) | <0.001 |
| Urban-rural residence MI | 0.103<br>(0.304) | 0.098<br>(0.297) | 0.103<br>(0.305) | 0.910 |
| Urban-residence MI | 0.062<br>(0.240) | 0.051<br>(0.221) | 0.009<br>(0.093) | 0.003 |
| New rural cooperative MI | 0.437<br>(0.496) | 0.636<br>(0.482) | 0.754<br>(0.431) | <0.001 |
| Governmental MI | 0.039<br>(0.193) | 0.014<br>(0.117) | 0.004<br>(0.066) | <0.001 |

*Notes:* P-values are calculated based on Chi-square test for categorical variables, and one-way ANOVA for continuous variables. Abbreviations: CI, cognitive impairment; SD, standard deviation; ADL, activities of daily living; IADL, instrumental activities of daily living; MI, medical insurance.

As illustrated in Figure 2 and 3, the performance of disease management is consistently lower among people in severer stages of CI, with the most salient difference in disease prevention. Overall, only 50% of the older adults have had physical examination within one year. Stratified by cognitive status, 52% of older adults with no CI have taken physical examination in the past year, followed by 45% among mild CI, and 43% among severe CI (Figure 2). In addition, 38% of the older adults have never take any physical examination within 3 year, and the difference between CI and non-CI is even larger, as compared to preventive behaviors within 1 year. Around 48% of persons with CI, either mild or severe CI, did not take any physical examination within 3 years. The percentage is comparatively lower among persons without CI, at 33%.

**Figure 2.**
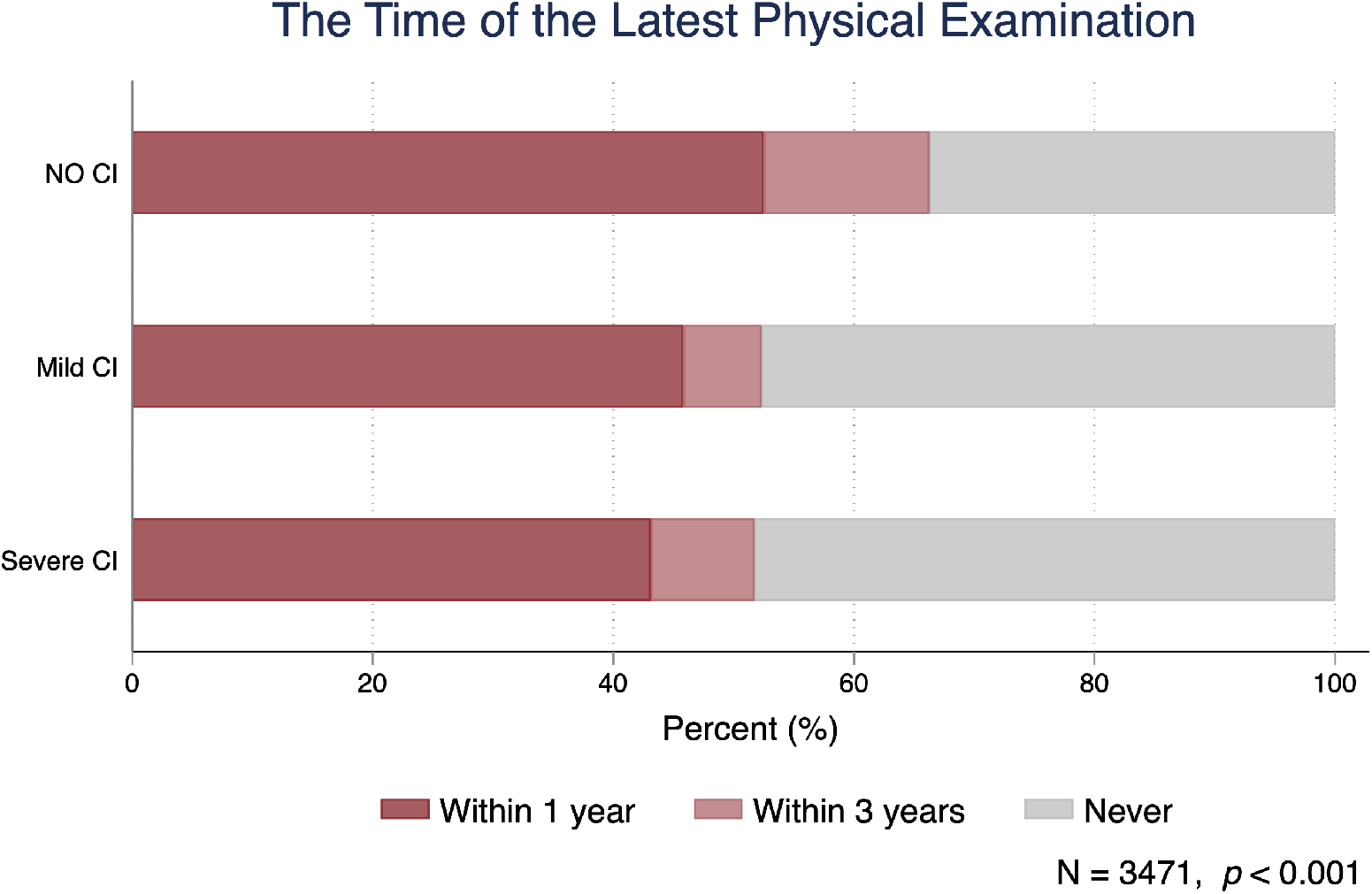
Preventive care utilization among older adults in different stages of cognitive impairment (CI) *Notes*: “Never” means that individuals have never had any physical examination in the past 3 years. P-value is calculated based on Chi-square test.

**Figure 3.**
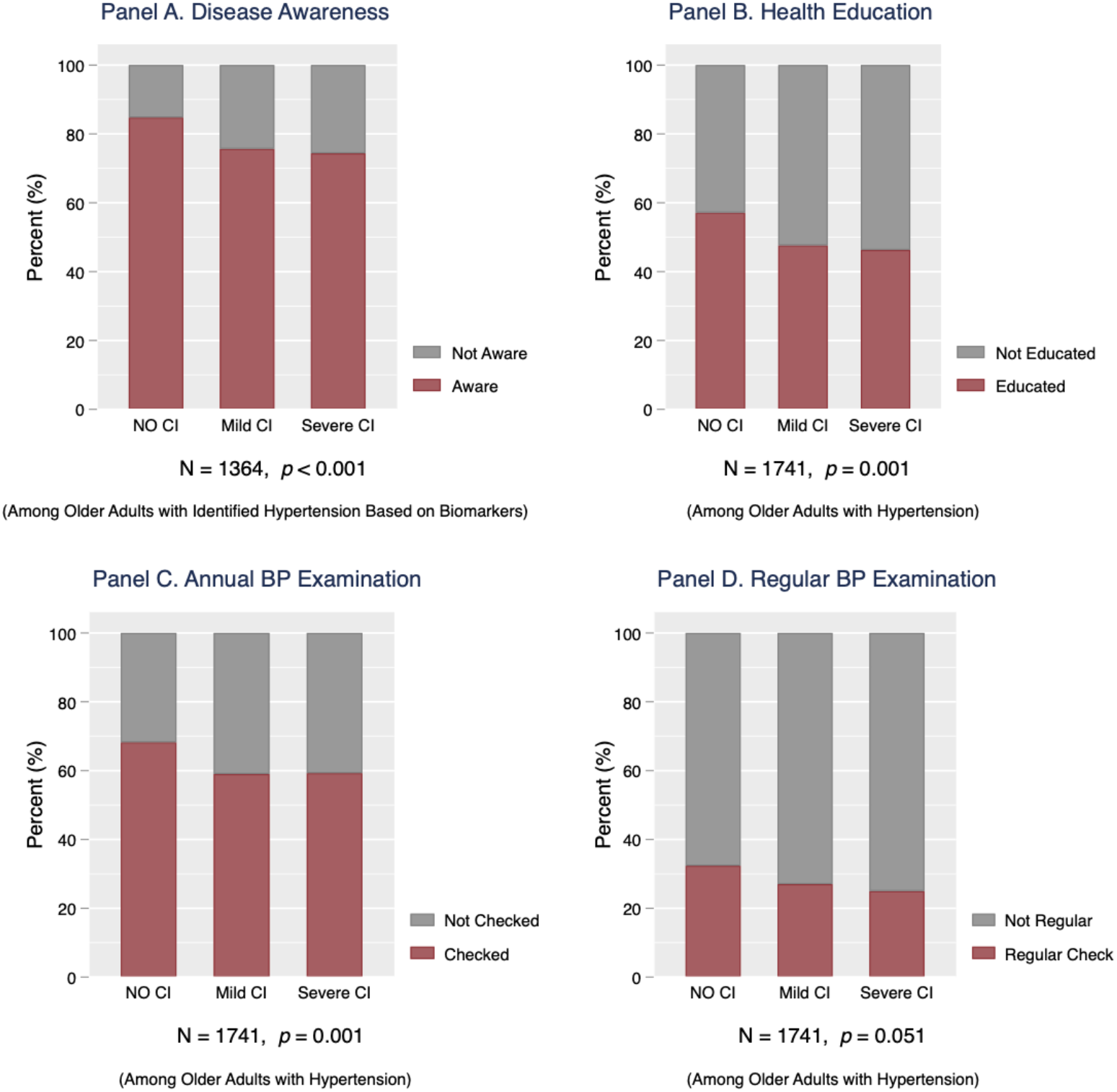
The management of hypertension among older adults in different stages of cognitive impairment (CI) *Notes*: Disease awareness denotes whether individuals with hypertension identified by biomarkers in the wave of 2015, have ever been diagnosed with hypertension by a doctor by the wave of 2018. Health education denotes whether individuals have ever been given health education/advice by doctors to control their hypertension, including weight control, exercise, diet and/or smoking control. Annual blood pressure (BP) examination denotes whether individuals have BP examination in the past year. Regular BP examination denotes whether individuals have had BP examination by community/village doctors regularly. Health education, annual BP examination, and regular BP examination are examined among individuals either have identified hypertension in the wave of 2015 or know they have hypertension as of the wave of 2018, i.e., with hypertension. P-values are calculated based on Chi-square test.

Similar pattern is observed for hypertension management (Figure 3). We show that for individuals with hypertension, those in severer stages of CI are less likely to be aware of hypertension, receive health education, and monitor their BP annually and regularly. Older adults overall have poor performance in hypertension management, with significantly lower performance among older adults with CI. The difference between individuals with and without CI is about 10% for each outcomes.

Furthermore, by examining access to LTSS, we show that older adults with CI lack both formal and informal supports (Figure 4)^5^. In particular, individuals in severer stages of CI tend to have less informal supports from spouse and friends. Although cognitively impaired adults have greater access to home-based and community-based services, the actual utilization rates are still quite low (i.e., less than 5%). These descriptive results point to the increased vulnerability of older adults with CI.

**Figure 4.**
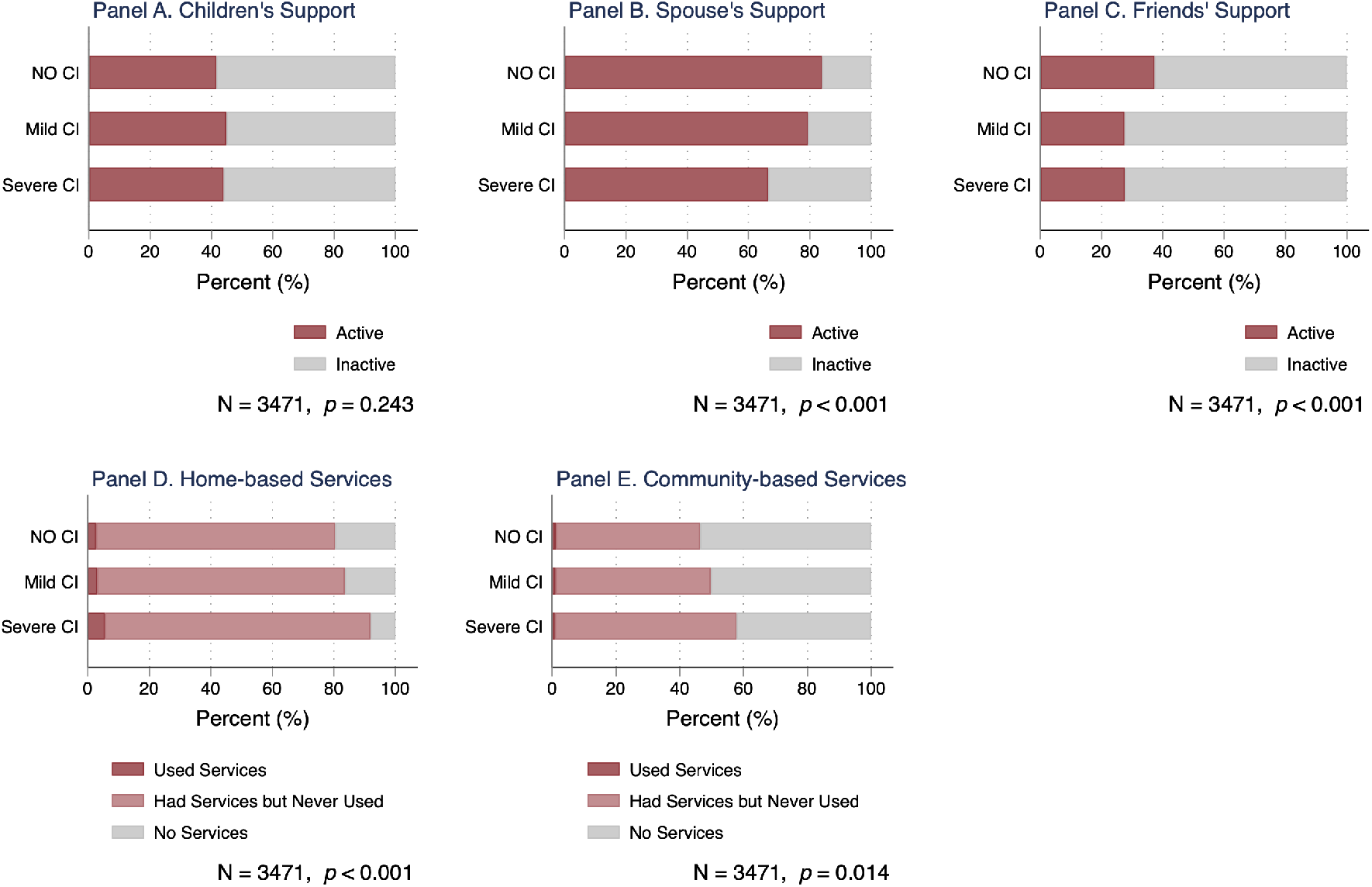
Access to various types of long-term services and supports (LTSS) among older adults in different stages of cognitive impairment (CI) *Notes*: Children’s support measures whether individuals have children’s visits more than median (active) or below (inactive). Spousal support measures whether individuals have spouse present (active) or not (inactive). Friends’ support measures whether individuals have interaction with friends in the last month (active) or not (inactive). Home-based services encompass family beds and onsite visit, and community-based services encompass day care centers and community nursing. For each service, access (i.e., having services or not) is measured at the local city level, whereas the utilization of service is assessed at the individual level. P-values are calculated based on Chi-square test.

### 3.2 Effects of LTSS and Cognitive Impairment on Disease Management

Our regression analysis corroborates the gradient relationship between CI and disease management. Relative to older adults with no CI, individuals with mild CI and severe CI have significantly lower odds of taking physical examination (Table 2), being aware of hypertension (Table 3), and getting annual and regular BP test (Table 4). Gaps are even greater for older adults with severe CI as compared to those with mild CI.

**Table 2.** Effects of cognitive impairment and LTSS on disease prevention

| VARIABLES | (1) | (2) |
| --- | --- | --- |
|  | Physical Examination in the Past Year |  |
| | $\beta$<br>(SE) | OR<br>[95% CI] |
| Mild CI | -0.281**<br>(0.102) | 0.75<br>[0.62 - 0.92] |
| Severe CI | -0.474*<br>(0.192) | 0.62<br>[0.43 - 0.91] |
| Children's Support | 0.055<br>(0.076) | 1.06<br>[0.91 - 1.23] |
| Spousal Support | 0.148<br>(0.108) | 1.16<br>[0.94 - 1.43] |
| Friends' Support | 0.272***<br>(0.079) | 1.31<br>[1.13 - 1.53] |
| Home-based Services | -0.121<br>(0.129) | 0.89<br>[0.69 - 1.14] |
| Community-based Services | -0.045<br>(0.095) | 0.96<br>[0.79 - 1.15] |
| Observations | 3,471 |  |
| Covariates | YES |  |
| Province FE | YES |  |
| Pseudo R-squared | 0.070 |  |

*Notes:* Logistic regression is conducted for disease prevention. Standard errors are clustered at city level. Preventive care utilization is assessed among all older adults. The regression model controls for age, gender, education, rural/urban status, number of chronic diseases, ADL, IADL, number of living children, and health insurance. Province fixed effects are included. Abbreviations: OR, odds ratio; SE, standard errors; CI, cognitive impairment; FE, fixed effects. Statistical significance: \*\*\* $p < 0.001$ , \*\* $p < 0.01$ , \* $p < 0.05$ .

**Table 3.**
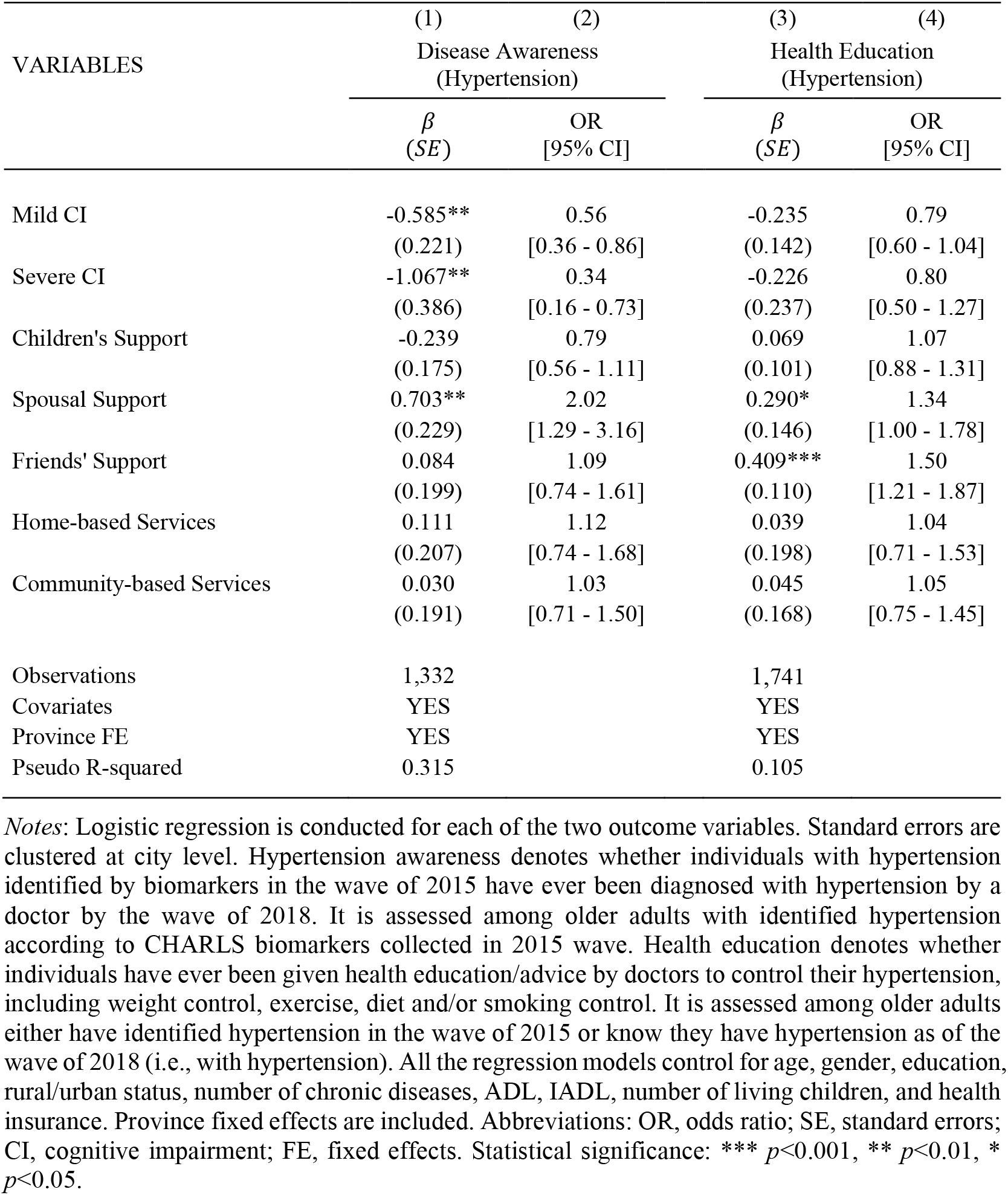
Effects of cognitive impairment and LTSS on the awareness and education of hypertension

| VARIABLES | (1) | (2) | (3) | (4) |
| --- | --- | --- | --- | --- |
|  | Disease Awareness<br>(Hypertension) |  | Health Education<br>(Hypertension) |  |
| | $\beta$<br>(SE) | OR<br>[95% CI] | $\beta$<br>(SE) | OR<br>[95% CI] |
| Mild CI | -0.585**<br>(0.221) | 0.56<br>[0.36 - 0.86] | -0.235<br>(0.142) | 0.79<br>[0.60 - 1.04] |
| Severe CI | -1.067**<br>(0.386) | 0.34<br>[0.16 - 0.73] | -0.226<br>(0.237) | 0.80<br>[0.50 - 1.27] |
| Children's Support | -0.239<br>(0.175) | 0.79<br>[0.56 - 1.11] | 0.069<br>(0.101) | 1.07<br>[0.88 - 1.31] |
| Spousal Support | 0.703**<br>(0.229) | 2.02<br>[1.29 - 3.16] | 0.290*<br>(0.146) | 1.34<br>[1.00 - 1.78] |
| Friends' Support | 0.084<br>(0.199) | 1.09<br>[0.74 - 1.61] | 0.409***<br>(0.110) | 1.50<br>[1.21 - 1.87] |
| Home-based Services | 0.111<br>(0.207) | 1.12<br>[0.74 - 1.68] | 0.039<br>(0.198) | 1.04<br>[0.71 - 1.53] |
| Community-based Services | 0.030<br>(0.191) | 1.03<br>[0.71 - 1.50] | 0.045<br>(0.168) | 1.05<br>[0.75 - 1.45] |
| Observations | 1,332 |  | 1,741 |  |
| Covariates | YES |  | YES |  |
| Province FE | YES |  | YES |  |
| Pseudo R-squared | 0.315 |  | 0.105 |  |

*Notes:* Logistic regression is conducted for each of the two outcome variables. Standard errors are clustered at city level. Hypertension awareness denotes whether individuals with hypertension identified by biomarkers in the wave of 2015 have ever been diagnosed with hypertension by a doctor by the wave of 2018. It is assessed among older adults with identified hypertension according to CHARLS biomarkers collected in 2015 wave. Health education denotes whether individuals have ever been given health education/advice by doctors to control their hypertension, including weight control, exercise, diet and/or smoking control. It is assessed among older adults either have identified hypertension in the wave of 2015 or know they have hypertension as of the wave of 2018 (i.e., with hypertension). All the regression models control for age, gender, education, rural/urban status, number of chronic diseases, ADL, IADL, number of living children, and health insurance. Province fixed effects are included. Abbreviations: OR, odds ratio; SE, standard errors; CI, cognitive impairment; FE, fixed effects. Statistical significance: \*\*\* $p < 0.001$ , \*\* $p < 0.01$ , \* $p < 0.05$ .

**Table 4.** Effects of cognitive impairment and LTSS on the monitoring of hypertension

| VARIABLES | (1) | (2) | (3) | (4) |
| --- | --- | --- | --- | --- |
|  | Annual BP Examination<br>(Hypertension) |  | Regular BP Examination<br>(Hypertension) |  |
| | $\beta$<br>(SE) | OR<br>[95% CI] | $\beta$<br>(SE) | OR<br>[95% CI] |
| Mild CI | -0.409**<br>(0.132) | 0.66<br>[0.51 - 0.86] | -0.419**<br>(0.153) | 0.66<br>[0.49 - 0.89] |
| Severe CI | -0.587*<br>(0.272) | 0.56<br>[0.33 - 0.95] | -0.836**<br>(0.283) | 0.43<br>[0.25 - 0.75] |
| Children's Support | 0.034<br>(0.111) | 1.03<br>[0.83 - 1.29] | -0.101<br>(0.115) | 0.90<br>[0.72 - 1.13] |
| Spousal Support | 0.489***<br>(0.143) | 1.63<br>[1.23 - 2.16] | 0.268<br>(0.140) | 1.31<br>[0.99 - 1.72] |
| Friends' Support | 0.397***<br>(0.112) | 1.49<br>[1.19 - 1.85] | 0.349**<br>(0.120) | 1.42<br>[1.12 - 1.79] |
| Home-based Services | 0.082<br>(0.148) | 1.09<br>[0.81 - 1.45] | 0.170<br>(0.180) | 1.19<br>[0.83 - 1.69] |
| Community-based Services | -0.039<br>(0.127) | 0.96<br>[0.75 - 1.23] | 0.312*<br>(0.136) | 1.37<br>[1.05 - 1.78] |
| Observations | 1,741 |  | 1,741 |  |
| Covariates | YES |  | YES |  |
| Province FE | YES |  | YES |  |
| Pseudo R-squared | 0.108 |  | 0.093 |  |

*Notes:* Logistic regression is conducted for each of the two outcome variables. Standard errors are clustered at city level. Annual BP examination denotes whether individuals have BP examination in the past year. Regular BP examination denotes whether individuals have had blood pressure examination by community/village doctors regularly. Both variables are assessed among older adults who either have identified hypertension in the wave of 2015 or know they have hypertension as of the wave of 2018 (i.e., with hypertension). All the regression models control for age, gender, education, rural/urban status, number of chronic diseases, ADL, IADL, number of living children, and health insurance. Province fixed effects are included. Abbreviations: OR, odds ratio; SE, standard errors; CI, cognitive impairment; FE, fixed effects. Statistical significance: \*\*\* $p < 0.001$ , \*\* $p < 0.01$ , \* $p < 0.05$ .

Adjusting for sociodemographic characteristics and geographical differences, significant protective effects of LTSS are found for a set of disease management outcomes. People with friends’ support are more likely to have physical examination in the past year [OR=1.31, 95%CI=1.13-1.53]. In contrast, we find no significant association of children’s support, spousal support, and home-based and community-based services with preventive care utilization (Table 2).

With regard to hypertension management, we show that spousal support has large and significant impact on hypertension awareness, health education and annual BP examination. In particular, as presented in Table 3 and Table 4, people with spouse present have 102% increased odds of being aware of hypertension, 34% increased odds of receiving health education, and 63% increased odds of doing annual BP examination than those without. On the other hand, Friends’ support has significant and positive effect on health education [OR=1.50, 95%CI=1.21-1.87], annual BP examination [OR=1.49, 95%CI=1.19 - 1.85] and regular BP examination [OR=1.42, 95%CI=1.12- 1.79]. Although home-based and community-based services generally have positive impact on hypertension management, we only observe significant association between community-based services and regular BP examination. In specific, older adults with local access to community-based services have 37% higher odds of getting regular BP examination than those without the access.

### 3.3 Effects of LTSS on Disease Management among Older adults with and without Cognitive Impairment

Since CI greatly worsens individuals’ ability to manage their diseases by themselves, the LTSS are potentially of crucial importance for cognitive impaired individuals. Meanwhile, CI can also create barriers for effectively seeking and utilizing the services, which can make it more difficult for cognitively impaired individuals to benefit from LTSS. To examine the possible heterogeneous effect of LTSS, we separately run the regressions for older adults with and without CI. The results are illustrated in Figure 5-7.

**Figure 5.**
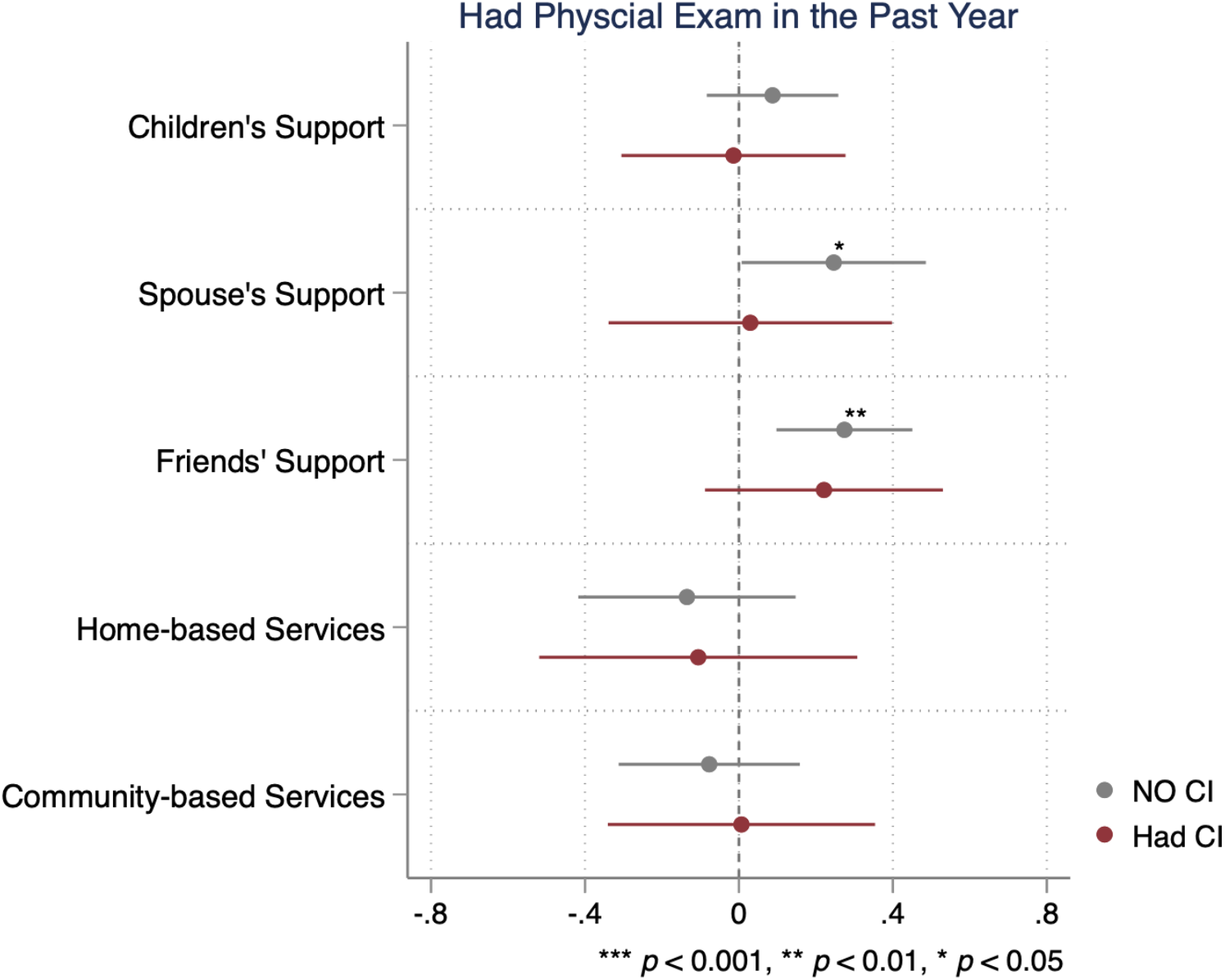
Effects of LTSS on disease prevention among older adults with and without cognitive impairment (CI) *Notes*: Logistic regressions are respectively conducted among sample without CI (shown in gray color; N = 2440) and sample with CI (shown in red color; N = 1030) to examine the association between LTSS and disease prevention. Plotted points represent the regression coefficients, and the horizontal lines represent the 95% confidence intervals. Standard errors are clustered at city level. All the regression models control for age, gender, education, rural/urban status, number of chronic diseases, ADL, IADL, number of living children, and health insurance. Province fixed effects are included. The detailed estimates, such as odds ratio, are presented in Appendix Table A1.

**Figure 6.**
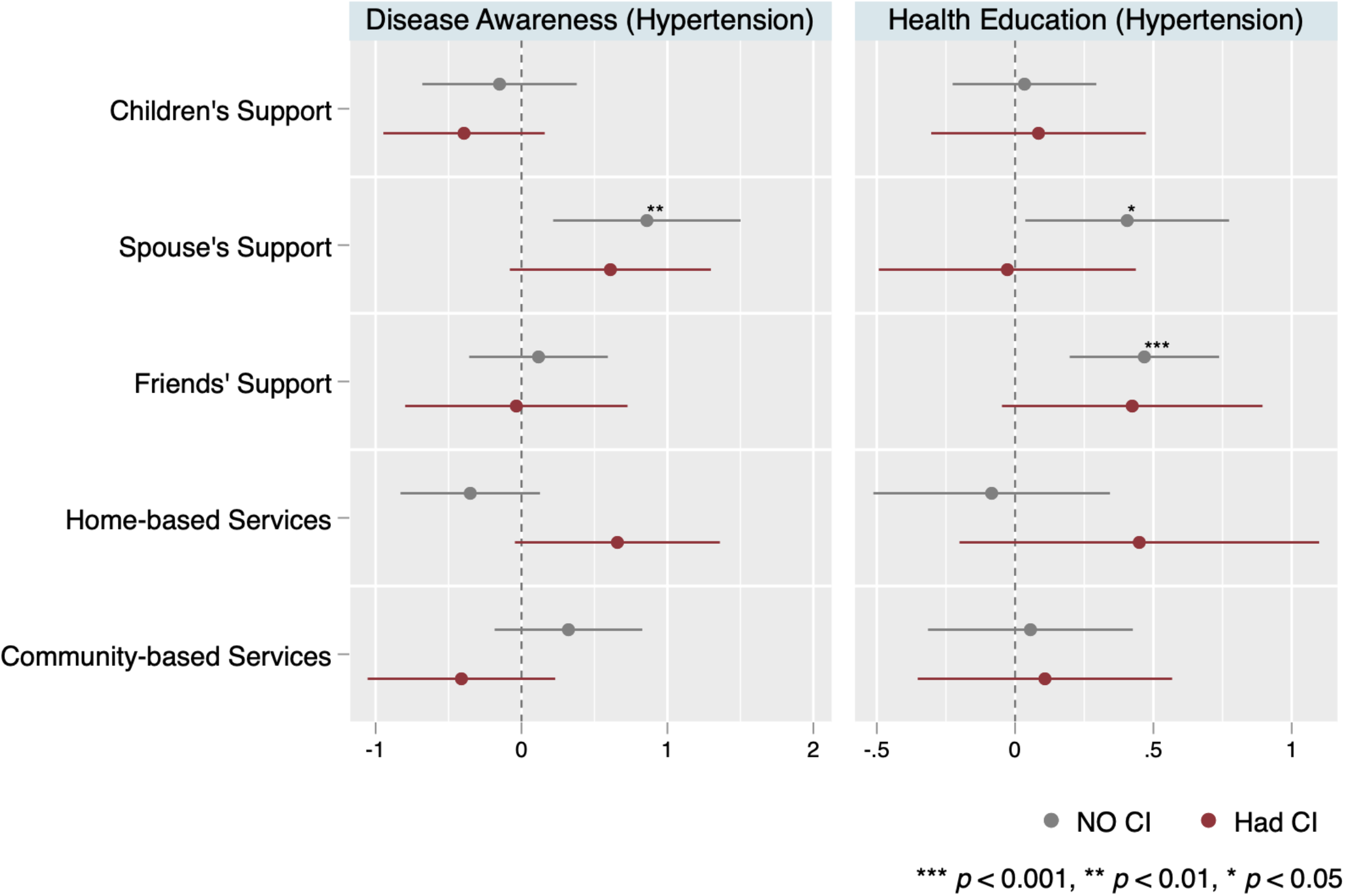
Effects of LTSS on the awareness and education of hypertension among older adults with and without cognitive impairment (CI) *Notes*: The left panel shows the logistic regression estimates for the awareness of hypertension, while the right panel shows the results for the health education of hypertension. Hypertension awareness denotes whether individuals with hypertension identified by biomarkers in the wave of 2015, have ever been diagnosed with hypertension by a doctor by the wave of 2018. It is assessed among older adults with identified hypertension according to the biomarkers of wave 2015. Health education denotes whether individuals have ever been given health education/advice by their doctors to control their hypertension, including weight control, exercise, diet and/or smoking control. It is assessed among older adults either have identified hypertension in the wave of 2015 or know they have hypertension as of the wave of 2018. Logistic regressions are respectively conducted among sample without CI (shown in gray color; N = 913 for awareness and N = 1211 for education) and sample with CI (shown in red color; N = 400 for awareness and N = 526 for education) to examine the association between LTSS and hypertension awareness and education. Plotted points represent the regression coefficients, and the horizontal lines represent the 95% confidence intervals. Standard errors are clustered at city level. All the regression models control for age, gender, education, rural/urban status, number of chronic diseases, ADL, IADL, number of living children, and health insurance. Province fixed effects are included. The detailed estimates, such as odds ratio, are presented in Appendix Table A2.

**Figure 7.**
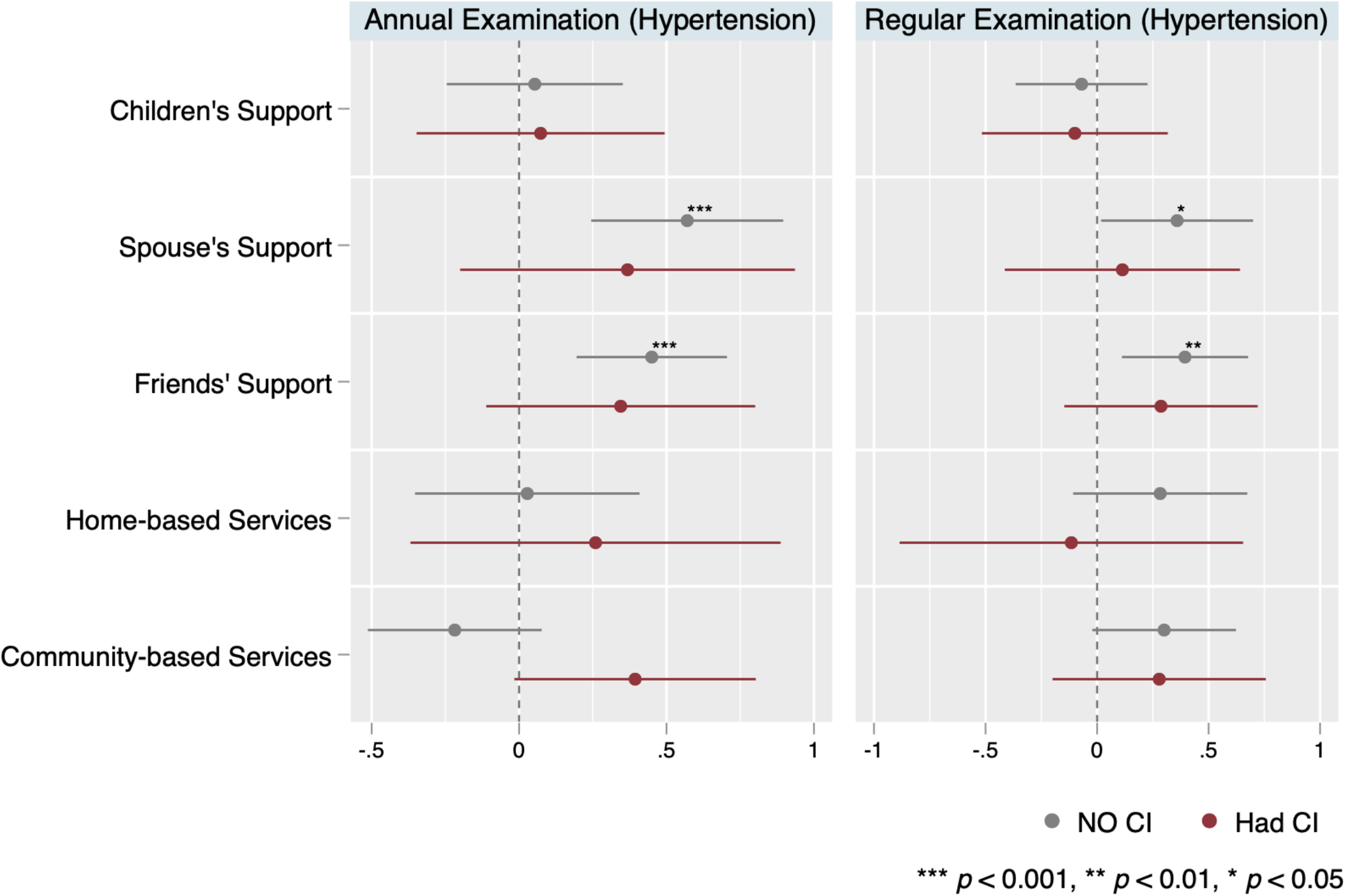
Effects of LTSS on the monitoring of hypertension among older adults with and without cognitive impairment (CI) *Notes*: The left panel shows the logistic regression estimates for annual blood pressure (BP) examination, while the right panel shows the results for regular BP examination. Annual BP examination denotes whether individuals have BP examination in the past year. Regular BP examination denotes whether individuals have had blood pressure examination by community/village doctors regularly. Both variables are assessed among older adults who either have identified hypertension in the wave of 2015 or know they have hypertension as of the wave of 2018 (i.e., with hypertension). Logistic regressions are respectively conducted among sample without CI (shown in gray color; N = 1191 annual BP examination and N=1205 for regular BP examination) and sample with CI (shown in red color; N = 522 for both variables) to examine the association between LTSS and hypertension monitoring. Plotted points represent the regression coefficients, and the horizontal lines represent the 95% confidence intervals. Standard errors are clustered at city level. All the regression models control for age, gender, education, rural/urban status, number of chronic diseases, ADL, IADL, number of living children, and health insurance. Province fixed effects are included. The detailed estimates, such as odds ratio, are presented in Appendix Table A3.

Figure 5 visualizes the results for disease prevention. Consistent with the baseline estimates, older adults with friends’ support are more likely to have physical examination in the past year. Nevertheless, we only observe large and significant effect among older adults without CI, while there is no significant association for cognitively impaired individuals. Notably, despite the overall effect of spousal support being insignificant, it has significant effect for older adults with no CI.

For hypertension management, the effects are generally larger and more significant for older adults without CI, while weaker for those with CI (Figure 6-7). In particular, spousal support only has positive and significant association with disease awareness, health education, annual BP examination and regular BP examination among cognitively intact individual, but not among impaired older adults. Similarly, friends’ support only significantly increases the odds of health education on hypertension, annual BP examination and regular BP examination among older adults with no CI. As for formal LTSS, although we do find some modest evidence that access to home-based services and community-based services respectively have larger positive effect on disease awareness and annual examination for individuals with CI than for those without CI, the effects of these formal services are merely significant at 10% level.

## 4. Discussion and Conclusion

Amid rapid population aging and rising burden of non-communicable diseases (NCDs), a sizable and still growing share of Chinese older adults live with mild CI or dementia. Consequently, the demand for services and supports will grow substantially, as effective disease prevention and control can be cognitively demanding. Using nationally representative physical examination and survey data from waves 2015 and 2018 of the China Health and Retirement Longitudinal Study (CHARLS), this paper is among the first to document the cognitive gradients in disease management and characterize the differential effects of long-term care services and supports (LTSS) on disease management among older adults by stage of CI. In particular, we examine the preventive care utilization and hypertension management, and highlight the potential challenges in supporting cognitively impaired older adults.

Several findings warrant further discussion. First of all, we find that older adults with severer stages of CI are facing higher vulnerability in disease management while receiving insufficient formal and informal supports. Compared to cognitively intact individuals, those with CI are about 15% less likely to utilize preventive care, and about 10% less likely to be aware of hypertension, receive health advice, or effectively monitor their BP. A lack of disease prevention and control will inevitably result in elevated disease risks overall, accelerating the progression of cognitive aging and widening the health gaps across the population. While LTSS may provide supports to these individuals, cognitively impaired adults are less likely to have spousal and friends’ support than those without CI. Moreover, the overall utilization rate of home-based and community-based services are quite low, even though they have access to these services. Therefore, given the weakening informal care under the changing demographic structure, more efforts should be devoted to improving the provision of formal long-term care and services and encouraging the effective utilization of the services, especially among cognitively impaired individuals.

Second, we demonstrate that LTSS play a critical role in facilitating disease management. In particular, informal LTSS, such as spousal support and friends’ support, can greatly increase preventive care use and help older adults manage and monitor their health and chronic conditions, such as hypertension. Although access to community-based services may slightly augment the probability of regular BP examination, home-based services generally have little impact on the overall performance of disease management. This is not surprising because home-based and community-based services in China are still underdeveloped and unequally distributed, despite recent efforts of Chinese government in promoting long-term care systems (Feng et al., 2020). Our findings re-emphasize the necessity and importance of promoting formal LTSS and establishing long-term care systems to provide accessible, adequate, and well-targeted supports for older adults, particularly given the limited and declining capacity of informal support (Feng et al., 2012).

Third, we show that individuals with CI are less likely to benefit from informal LTSS than those without CI. The effects of spousal support and friend’s support on disease management are consistently lower and weaker among older adults with CI as compared to those without CI. This pattern is broadly observed for preventive care utilization as well as hypertension awareness, health education, and monitoring, which implies the challenges cognitively impaired older adults face in benefiting from informal care. Older adults with CI or dementia often need comprehensive and continuous support from caregivers; however, the co-occurred behavioral and psychological symptoms can make it extremely hard for informal caregivers to understand their actual demands and feelings, thereby imposing large barriers for caregivers to offering appropriate care and support (Savva et al., 2009). Publicly-funded training may be provided to help family and other informal caregivers possess professional skills and knowledge (Wang et al., 2018). Yet, with the increasingly strained manpower of informal caregiving, formal home care or community services are urgently needed. With regard to the current home and community-based services, we do find some modest evidence that these services may somewhat equalize the differences in disease management between people with and without CI. Nonetheless, the effect is too small to be significant. There is still a long way to go for the Chinese formal long-term care system to be established and developed.

Our analysis has some limitations and weaknesses. First, we rely on survey data to investigate LTSS, which is subject to recall biases and inaccuracies. It may be of interest for future studies to use administrative data to more accurately measure the availability of LTSS and evaluate their impacts. Second, the measures of informal care may not fully reflect the actual supports individuals received. CHARLS did not collect adequately rich information on informal caregiving for all participants, so we are unable to directly assess the intensity of support individuals receive. Third, as the MMSE and biomarkers are not available in the same wave, we use two waves of survey to construct the measures of hypertension management. Although it may somewhat overestimate the overall performance among older adults, it allows us to identify those who have more issues with the health awareness and control since we give participants a longer period (3-year more) to be aware of their conditions. Our results thus may imply a more salient difference in disease management compared to the estimates using the information of a single wave^6^. Fourth, only cross-sectional data of MMSE are collected, which do not allow us to examine how the transition from no-CI to mild-CI or severe-CI would interfere with the preventive care utilization and chronic disease management. Follow-up surveys are needed to understand the temporal relationship. Finally, due to the limited sample size, it is infeasible to further stratify our sample by rural/urban, education level, or other sociodemographic conditions. Future studies may combine multiple survey data to understand the heterogeneity across various social groups.

Despite these limitations, this study provides novel evidence on the impact of LTSS and CI on disease management, and reveals the difficulties and challenges for older adults with CI to benefit from LTSS. Our findings may inspire future research to identify the exact barriers in existing practice as well as the effective strategies for supporting vulnerable populations.

## Data Availability

Data are obtained from China Health and Retirement Longitudinal Study (CHARLS), a nationally representative survey of Chinese older adults age 45 years or older. The national baseline was conducted in 2011, and the respondents were longitudinally followed up in 2013, 2015, and 2018. All waves are publicly available upon submitting a form of data user agreement.

http://charls.pku.edu.cn/index/en.html

## Appendix

**Figure A1.**
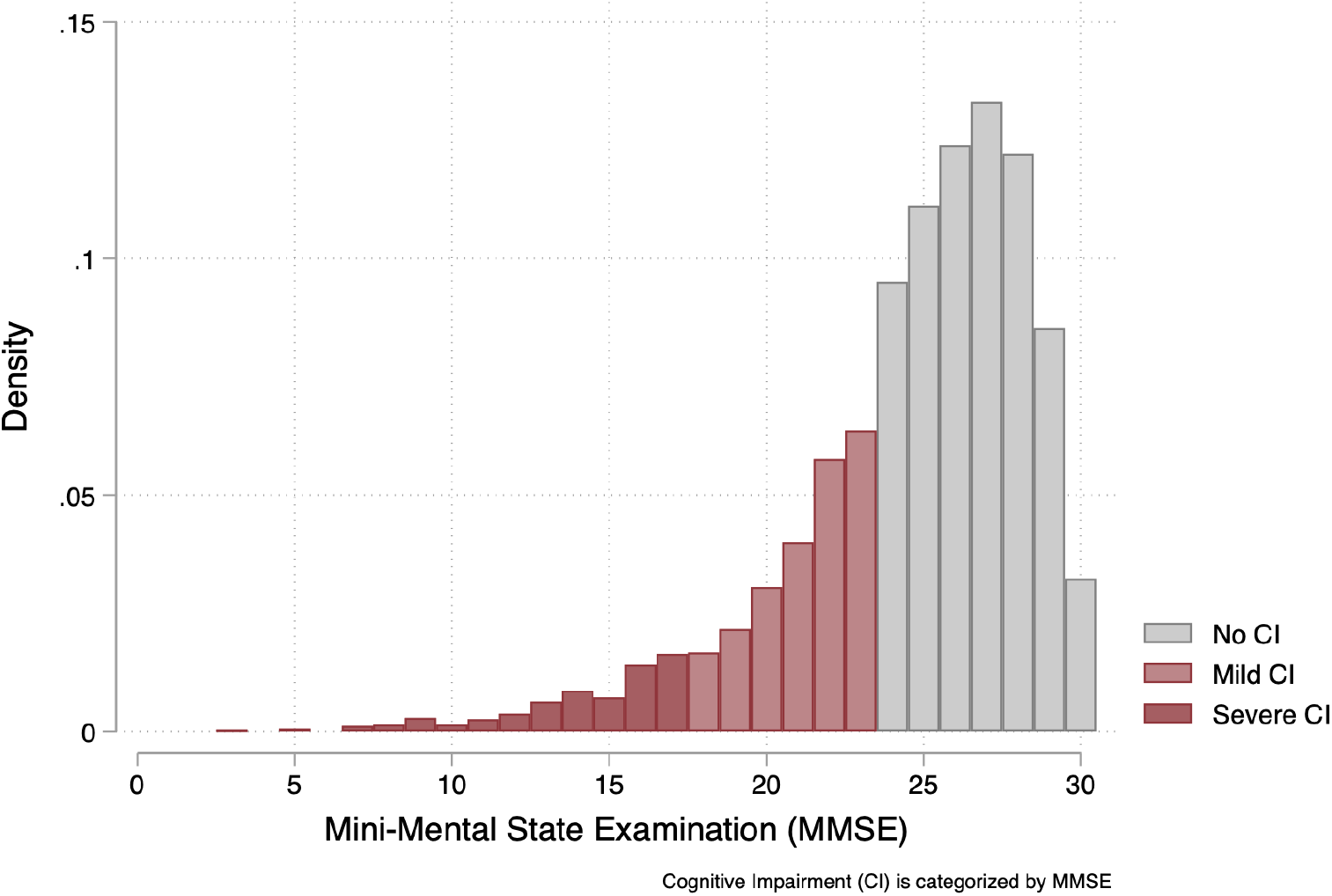
Distribution of Mini-Mental State Examination (MMSE) and different stages of cognitive impairment (CI) *Notes*: Cognitive impairment (CI) is categorized by the score of MMSE (0-30), with a score of 24-30 as “No CI”, a score of 18-23 as “Mild CI”, and a score of 0-17 as “Severe CI”. In our study sample (N=3471), the mean value of MMSE is 24.59; the standard deviation is 3.98; the minimum value is 3; and the maximum value is 30.

**Figure A2.**
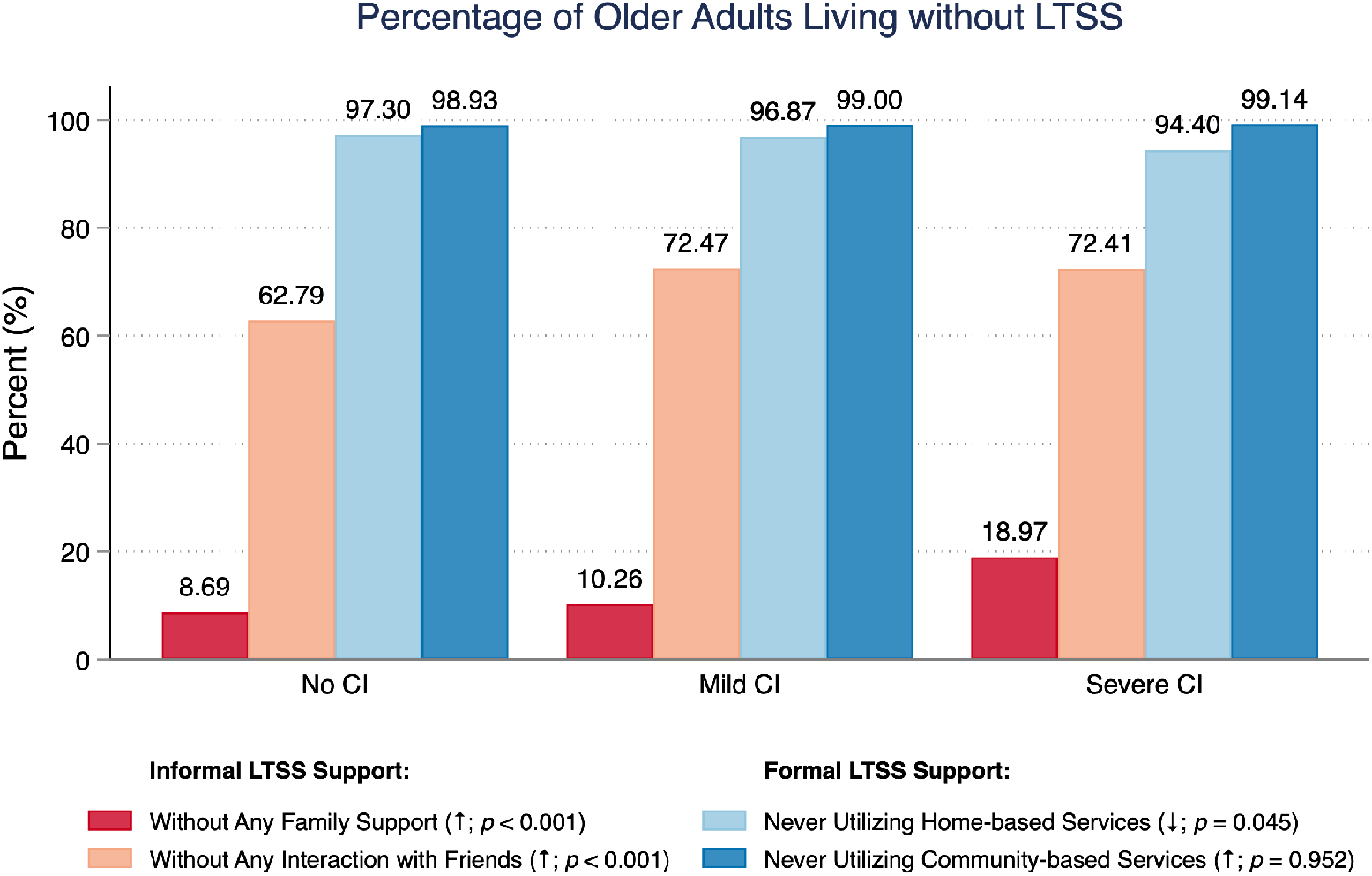
Percentage of older adults living without informal or formal long-term services and supports (LTSS) *Notes*: Family support and interaction with friends are used to represent informal LTSS, and home-based and community-based services are used to represent formal LTSS. Among them, family support measures whether individuals have either active children’s support (i.e., frequent children’s visits) or spousal support (i.e., spouse present). Home-based services encompass family beds and onsite visit, and community-based services encompass day care centers and community nursing. For better illustration, we only show the percentage of individuals who have never utilized formal services assessed at the individual level, while the percentage of individuals who have no access to formal services in local city is not presented. An upward arrow in the parentheses denotes that the percentage is higher for individuals with severer cognitive impairment, whereas a downward arrow denotes that the parentage is lower for individuals with severer cognitive impairment. P-values are calculated based on Chi-square test.

**Table A1.** Effects of LTSS on disease prevention among older adults with and without cognitive impairment (CI)

| VARIABLES | (1) | (2) | (3) | (4) |
| --- | --- | --- | --- | --- |
|  | Physical Examination in the Past Year<br>(No CI) |  | Physical Examination in the Past Year<br>(Had CI) |  |
| | $\beta$<br>(SE) | OR<br>[95% CI] | $\beta$<br>(SE) | OR<br>[95% CI] |
| Children's Support | 0.087<br>(0.087) | 1.09<br>[0.92 - 1.29] | -0.014<br>(0.149) | 0.99<br>[0.74 - 1.32] |
| Spousal Support | 0.246*<br>(0.122) | 1.28<br>[1.01 - 1.63] | 0.030<br>(0.188) | 1.03<br>[0.71 - 1.49] |
| Friends' Support | 0.274**<br>(0.090) | 1.32<br>[1.10 - 1.57] | 0.221<br>(0.158) | 1.25<br>[0.92 - 1.70] |
| Home-based Services | -0.135<br>(0.144) | 0.87<br>[0.66 - 1.16] | -0.106<br>(0.211) | 0.90<br>[0.60 - 1.36] |
| Community-based Services | -0.077<br>(0.120) | 0.93<br>[0.73 - 1.17] | 0.007<br>(0.177) | 1.01<br>[0.71 - 1.42] |
| Observations | 2,440 |  | 1,030 |  |
| Covariates | YES |  | YES |  |
| Province FE | YES |  | YES |  |
| Pseudo R-squared | 0.066 |  | 0.099 |  |

*Notes:* Logistic regressions are respectively conducted among sample without CI (column 1, 2) and sample with CI (column 3, 4). Standard errors are clustered at city level. All the regression models control for age, gender, education, rural/urban status, number of chronic diseases, ADL, IADL, number of living children, and health insurance. Province fixed effects are included. Abbreviations: OR, odds ratio; SE, standard errors; CI, cognitive impairment; FE, fixed effects. Statistical significance: \*\*\* $p < 0.001$ , \*\* $p < 0.01$ , \* $p < 0.05$ .

**Table A2.** Effects of LTSS on the awareness and education of hypertension among older adults with and without cognitive impairment

| VARIABLES | (1) | (2) | (3) | (4) | (5) | (6) | (7) | (8) |
| --- | --- | --- | --- | --- | --- | --- | --- | --- |
|  | Hypertension Awareness<br>(No CI) |  | Hypertension Awareness<br>(Had CI) |  | Hypertension Education<br>(No CI) |  | Hypertension Education<br>(Had CI) |  |
| | $\beta$<br>(SE) | OR<br>[95% CI] | $\beta$<br>(SE) | OR<br>[95% CI] | $\beta$<br>(SE) | OR<br>[95% CI] | $\beta$<br>(SE) | OR<br>[95% CI] |
| Children's Support | -0.151<br>(0.270) | 0.86<br>[0.51 - 1.46] | -0.394<br>(0.282) | 0.67<br>[0.39 - 1.17] | 0.034<br>(0.133) | 1.03<br>[0.80 - 1.34] | 0.085<br>(0.198) | 1.09<br>[0.74 - 1.60] |
| Spousal Support | 0.859**<br>(0.328) | 2.36<br>[1.24 - 4.49] | 0.609<br>(0.352) | 1.84<br>[0.92 - 3.66] | 0.405*<br>(0.188) | 1.50<br>[1.04 - 2.17] | -0.028<br>(0.237) | 0.97<br>[0.61 - 1.55] |
| Friends' Support | 0.116<br>(0.243) | 1.12<br>[0.70 - 1.81] | -0.036<br>(0.389) | 0.97<br>[0.45 - 2.07] | 0.467***<br>(0.138) | 1.60<br>[1.22 - 2.09] | 0.423<br>(0.240) | 1.53<br>[0.95 - 2.45] |
| Home-based Services | -0.351<br>(0.244) | 0.70<br>[0.44 - 1.14] | 0.657<br>(0.358) | 1.93<br>[0.96 - 3.89] | -0.085<br>(0.218) | 0.92<br>[0.60 - 1.41] | 0.449<br>(0.332) | 1.57<br>[0.82 - 3.00] |
| Community-based Services | 0.322<br>(0.258) | 1.38<br>[0.83 - 2.29] | -0.412<br>(0.328) | 0.66<br>[0.35 - 1.26] | 0.055<br>(0.189) | 1.06<br>[0.73 - 1.53] | 0.108<br>(0.235) | 1.11<br>[0.70 - 1.76] |
| Observations | 913 |  | 400 |  | 1,211 |  | 526 |  |
| Covariates | YES |  | YES |  | YES |  | YES |  |
| Province FE | YES |  | YES |  | YES |  | YES |  |
| Pseudo R-squared | 0.363 |  | 0.331 |  | 0.127 |  | 0.138 |  |

*Notes:* Logistic regressions are respectively conducted among sample without CI (column 1, 2, 5, 6) and sample with CI (column 3, 4, 7, 8) for each outcome variable. Hypertension awareness denotes whether individuals with hypertension identified by biomarkers in the wave of 2015 have ever been diagnosed with hypertension by a doctor by the wave of 2018. It is assessed among older adults with identified hypertension according to the biomarkers of wave 2015. Hypertension education denotes whether individuals have ever been given health education/advice by doctors to control their hypertension, including weight control, exercise, diet and/or smoking control. It is assessed among older adults either have identified hypertension in the wave of 2015 or know they have hypertension as of the wave of 2018. Standard errors are clustered at city level. All the regression models control for age, gender, education, rural/urban status, number of chronic diseases, ADL, IADL, number of living children, and health insurance. Province fixed effects are included. Abbreviations: OR, odds ratio; SE, standard errors; FE, fixed effects. Statistical significance: \*\*\* $p < 0.001$ , \*\* $p < 0.01$ , \* $p < 0.05$ .

**Table A3.** Effects of LTSS on the monitoring of hypertension among older adults with and without cognitive impairment (CI)

| VARIABLES | (1) | (2) | (3) | (4) | (5) | (6) | (7) | (8) |
| --- | --- | --- | --- | --- | --- | --- | --- | --- |
|  | Annual BP Examination<br>(No CI) |  | Annual BP Examination<br>(Had CI) |  | Regular BP Examination<br>(No CI) |  | Regular BP Examination<br>(Had CI) |  |
| | $\beta$<br>(SE) | OR<br>[95% CI] | $\beta$<br>(SE) | OR<br>[95% CI] | $\beta$<br>(SE) | OR<br>[95% CI] | $\beta$<br>(SE) | OR<br>[95% CI] |
| Children's Support | 0.053<br>(0.152) | 1.05<br>[0.78 - 1.42] | 0.073<br>(0.215) | 1.08<br>[0.71 - 1.64] | -0.069<br>(0.151) | 0.93<br>[0.69 - 1.25] | -0.100<br>(0.213) | 0.91<br>[0.60 - 1.37] |
| Spousal Support | 0.570***<br>(0.166) | 1.77<br>[1.28 - 2.45] | 0.368<br>(0.290) | 1.44<br>[0.82 - 2.55] | 0.359*<br>(0.174) | 1.43<br>[1.02 - 2.01] | 0.114<br>(0.269) | 1.12<br>[0.66 - 1.90] |
| Friends' Support | 0.450***<br>(0.130) | 1.57<br>[1.22 - 2.02] | 0.345<br>(0.233) | 1.41<br>[0.89 - 2.23] | 0.394**<br>(0.144) | 1.48<br>[1.12 - 1.97] | 0.287<br>(0.221) | 1.33<br>[0.86 - 2.06] |
| Home-based Services | 0.028<br>(0.194) | 1.03<br>[0.70 - 1.50] | 0.259<br>(0.320) | 1.30<br>[0.69 - 2.43] | 0.283<br>(0.199) | 1.33<br>[0.90 - 1.96] | -0.115<br>(0.393) | 0.89<br>[0.41 - 1.93] |
| Community-based Services | -0.218<br>(0.150) | 0.80<br>[0.60 - 1.08] | 0.394<br>(0.209) | 1.48<br>[0.98 - 2.23] | 0.301<br>(0.164) | 1.35<br>[0.98 - 1.86] | 0.279<br>(0.244) | 1.32<br>[0.82 - 2.13] |
| Observations | 1,191 |  | 522 |  | 1,205 |  | 522 |  |
| Covariates | YES |  | YES |  | YES |  | YES |  |
| Province FE | YES |  | YES |  | YES |  | YES |  |
| Pseudo R-squared | 0.122 |  | 0.130 |  | 0.124 |  | 0.094 |  |

*Notes:* Logistic regressions are respectively conducted among sample without CI (column 1, 2, 5, 6) and sample with CI (column 3, 4, 7, 8) for each outcome variable. Annual BP examination denotes whether individuals have BP examination in the past year. Regular BP examination denotes whether individuals have had blood pressure examination by community/village doctors regularly. Both variables are assessed among older adults who either have identified hypertension in the wave of 2015 or know they have hypertension as of the wave of 2018 (i.e., with hypertension). All the regression models control for age, gender, education, rural/urban status, number of chronic diseases, ADL, IADL, number of living children, and health insurance. Province fixed effects are included. Abbreviations: OR, odds ratio; SE, standard errors; CI, cognitive impairment; FE, fixed effects. Statistical significance: \*\*\* $p < 0.001$ , \*\* $p < 0.01$ , \* $p < 0.05$ .

## Footnotes

1 As the explanatory variables (i.e., cognition and LTSS) are only available in 2018, we evaluate the hypertension diagnosis/awareness and management also using the self-reported information collected in 2018.

2 In our sample, only a small proportion of people have ever received these services.

3 To avoid self-selection problem, we measure whether individuals had access to home and community-based services in the local city rather than individuals’ utilization of the services themselves.

4 We choose not to include household income and wealth due to a large number of missingness in income and asset variables, which can greatly reduce statistical power given the limited sample size in our study. Education level, rural/urban status are instead used as proxy measures of socioeconomic conditions.

5 See appendix Figure A2 for more comparisons.

6 In fact, even if we use instant measures of awareness, the estimates may not improve since there can be more measurement error given the instant timing and delayed learning effect.

